# Cumulative Effects of Resting-state Connectivity Across All Brain Networks Significantly Correlate with ADHD Symptoms

**DOI:** 10.1101/2021.11.16.21266121

**Authors:** Michael A. Mooney, Robert J.M. Hermosillo, Eric Feczko, Oscar Miranda-Dominguez, Lucille A. Moore, Anders Perrone, Nora Byington, Gracie Grimsrud, Amanda Rueter, Elizabeth Nousen, Dylan Antovich, Sarah W. Feldstein Ewing, Bonnie J. Nagel, Joel T. Nigg, Damien A. Fair

## Abstract

**Background:** The clinical utility of MRI neuroimaging studies of psychopathology has been limited by a constellation of factors—small sample sizes, small effect sizes, and heterogeneity of methods and samples across studies—that hinder generalizability and specific replication. An analogy is early genomics studies of complex traits, wherein a move to large, multi-site samples and a focus on cumulative effects (polygenic scores) led to reproducible and clinically applicable effects from genome-wide association studies. A similar logic in MRI may provide a way to improve reproducibility, precision, and clinical utility for brain-wide MRI association studies.

**Methods:** Polyneuro scores (PNS) represent the cumulative effect of brain-wide measures—in the present case, resting-state functional connectivity (rs-fcMRI) associated with ADHD symptoms. These scores were constructed and validated using baseline data from the Adolescent Brain Cognitive Development (ABCD, N=5666) study, with a reproducible matched subset as the discovery cohort (N=2801). Association between the PNS and ADHD symptoms was further tested in an independent case-control cohort, the Oregon-ADHD-1000 (N=533).

**Results:** The ADHD PNS was significantly associated with ADHD symptoms in both the ABCD and Oregon cohorts after accounting for relevant covariates (p-values < 0.001). While the strongest effects contributing to the PNS were concentrated among connections involving the default mode and cingulo-opercular networks, the most predictive PNS involved connectivity across all brain networks. These findings were robust to stringent motion thresholds. In the longitudinal Oregon-ADHD-1000, non-ADHD comparison youth had significantly lower ADHD PNS (β=-0.309, p=0.00142) than children with persistent ADHD (met diagnostic criteria at two or more time points from age 7 to 19). The ADHD PNS, however, did not reliably mediate polygenic risk for ADHD. Instead, the PNS and an ADHD polygenic score were independently associated with ADHD symptoms.

**Conclusions:** A polyneuro risk score representing cumulative ADHD-associated resting-state connectivity was robustly associated with ADHD symptoms in two independent cohorts using distinct sampling designs, yet was independent of polygenic liability for ADHD, suggesting the need to examine environmental influences. The polyneuro score approach holds promise for improving the reproducibility of neuroimaging studies, identifying their clinical utility, and unraveling the complex relationships between brain connectivity and the etiology of behavioral disorders.

## INTRODUCTION

Attention-deficit hyperactivity disorder (ADHD) is a common neurodevelopmental disorder, affecting approximately 5% of school-aged children (Song, Dieckmann, and Nigg 2019; Cordova et al. 2021) and is a serious condition associated with future depression, substance use, antisocial behavior, under-employment, and shortened lifespan due to accidental death, suicide, and physical health complications. ADHD as a syndrome is characterized by age-inappropriate cognitive, behavioral, and emotional problems, such as inattention, hyperactivity/impulsivity, irritability, and anxiety, though there is significant heterogeneity in both presentation and outcome (Sonuga-Barke 2005; Posner, Park, and Wang 2014; Luo et al. 2019; Nigg, Karalunas, Feczko, et al. 2020; Fair et al. 2012). Symptoms typically develop in childhood, but can persist into adulthood for a significant proportion of those affected (Sibley et al. 2017). Substantial behavioral and quantitative genetic evidence suggests that ADHD symptoms exist in the population along a continuum and so can be meaningfully studied as a dimensional trait (as we do here) (Thapar 2018).

Neuroimaging studies have aimed to elucidate the brain mechanisms and correlates of ADHD as both a category and a symptom dimension. Although these investigations have identified a variety of distinct and localized structural and functional brain features associated with ADHD (Albajara Sáenz, Villemonteix, and Massat 2019; Hoogman et al. 2020; Mooney et al. 2021), results have not yet yielded reliable replication. This is likely secondary to relatively small brain-behavior effect sizes (Pereira-Sanchez and Castellanos 2021; Feczko et al. 2021; Marek et al. 2020; Smith and Nichols 2018). Nonetheless, considerable evidence supports the proposal that biological and physiological correlates of ADHD are not simply localized alterations in brain function, but rather constitute widely distributed functional brain systems as evaluated by resting state functional connectivity MRI (rs-fcMRI) studies. For example, a systematic review of rs-fcMRI studies of ADHD through 2013 found 26 studies with the number of ADHD cases analyzed ranging from 16 to 648 (Posner, Park, and Wang 2014). Although there were some consistent findings, such as altered connectivity within the default mode network (DMN) (Castellanos et al. 2008; Fair et al. 2010; Qiu et al. 2011), an overall conclusion was that rs-fcMRI studies do not support the hypothesis of a single neurocognitive deficit in ADHD. Rather, study results suggest that multiple brain networks are likely involved, providing support of a neurobiological basis for the heterogeneity observed in ADHD presentation (Saad, Griffiths, and Korgaonkar 2020).

More recently, meta-analyses of rs-fcMRI studies have provided further evidence that connectivity in multiple brain networks is associated with ADHD. However, these studies have primarily focused on a few hypothesized networks, rather than examining effects brain-wide. While these networks appear to participate in ADHD, they likely are not the full sum of brain involvement. In addition, the rapid increase in available data and efforts at data pooling are hampered by the heterogeneity of studies meta-analyzed (differences in the populations studied, data collection, and analysis methods), often leading to inconsistent findings. We highlight key findings in the literature for context.

Gao *et al.* examined results from 21 rs-fcMRI studies with a total of 700 ADHD cases and 580 controls, focusing on DMN, frontoparietal (FPN), and affective network (AN) connectivity (Gao et al. 2019). Hyperconnectivity between the FPN and regions of the DMN and AN, as well as hyperconnectivity between the FPN and regions of the ventral attention network (VAN) and the somatosensory network (SSN) were significantly associated with ADHD. Sutcubasi *et al.* meta-analyzed 20 seed-based rs-fcMRI studies that focused on four networks: DMN, cognitive control, salience, and affective/motivational (Sutcubasi et al. 2020). The studies comprised 944 ADHD cases and 1121 controls. Analyses restricted to children and adolescents found reduced connectivity between the DMN and all three other networks examined. Thus, results from these recent meta-analyses suggest that features of ADHD are associated with aberrant connectivity both within DMN and between DMN and cognitive control networks, but that other networks are involved as well.

Most striking, however, was that in a large meta-analysis (1094 ADHD cases, 884 controls) of both seed-based (18 studies) and non-seed-based (12 studies) whole-brain rs-fcMRI studies, Cortese *et al.* found no significant spatial convergence of ADHD-associated connectivity patterns (Cortese et al. 2021). The authors note that given the differences in meta-analytic methods and the criteria for study selection (hypothesis-free, compared to the focus on a specific subset of networks in previous meta-analyses), discordant results were not surprising. They offer two hypotheses to explain the conflicting findings: (a) that dysconnectivity patterns (i.e., the networks involved) differ among individuals with ADHD, or (b) that the precise location of the ADHD-associated network dysconnectivity differs among affected individuals. Both seem plausible given the heterogeneity among ADHD patients and current ADHD neuroimaging findings. A third hypothesis, consistent with the former, is the idea that ADHD-associated dysconnectivity is more broadly distributed across the entire brain, consisting of many small effects, and that it is the cumulative effect that correlates best with behavior.

These reviews highlight multiple linked challenges to brain-behavior associations with ADHD: (1) most brain-behavior associations for individual connections and/or brain regions show small effect sizes (Smith and Nichols 2018; Marek et al. 2020), (2) most studies use relatively small sample sizes underpowered to detect small effects (Posner, Park, and Wang 2014; Marek et al. 2020), (3) differences among study samples and methods continue to represent challenges for reproducibility (Marek et al. 2020), (4) ADHD-related connectivity or dysconnectivity may show inter-individual differences (Feczko et al. 2019; Nigg, Karalunas, Feczko, et al. 2020; Feczko and Fair 2020), and (5) recent evidence suggests the involvement of multiple brain networks in ADHD. Collectively, these challenges point toward the importance and value of taking a whole-brain perspective when studying the neural correlates of ADHD.

For instance, measures that represent the cumulative trait-associated effects of task-based fMRI measures across the entire brain were recently reported to explain a significantly greater proportion of trait variance than individual voxels (Zhao et al. 2019). In that study, a brain-wide association analysis was performed to identify voxels associated with performance on specific cognitive tasks. Those association measures were then used to construct a weighted sum of activity across the entire brain, which constitutes a single fMRI-based summary measure of the trait. Here we build on this concept by applying it to rs-fcMRI data, and generating scores representing brain-wide ADHD-associated connectivity.

Brain-wide summary scores, which we refer to as *polyneuro* scores (PNS; akin to polyvertex scores (Zhao et al. 2019)), are somewhat analogous to polygenic scores (Choi, Mak, and O’Reilly 2020; Ronald, Bode, and Polderman 2021; Li and He 2021) that represent cumulative genetic risk across the genome. PNS address a number of limitations of neuroimaging studies, similar to the way polygenic scores (PRS) addressed the challenges of early genomic studies.

First, PNS take advantage of large-scale consortium-level data sets [like those from the Adolescent Brain Cognitive Development (ABCD) study] (Feczko et al. 2021; Casey et al. 2018; Miller et al. 2016) to generate brain-wide effect estimates, and allow testing of cumulative effects in smaller data sets that would be underpowered to detect small effects shown in meta-analyses (Marek et al. 2020; Pereira-Sanchez and Castellanos 2021). In other words, it may not be possible to conduct a reproducible brain-wide association analysis in a small data set (e.g., N=200), but it would be possible to test the association between a PNS (built based on results from a large, independent discovery data set of N>2000) and the trait of interest. Second, PNS allow for heterogeneity among study subjects, because each individual gets a single summary score and the same cumulative effect can result from varying effects of individual brain features. And third, cumulative brain-wide effects should provide significantly greater predictive power than individual brain features, if the trait of interest truly has signal distributed across the brain.

The increased predictive power (effect size) of the PNS will also enable the examination of mediation effects, providing insight into the mechanisms of genetic or early environmental risk factors. Previous work has provided evidence for both structural and functional MRI measures partially mediating the effect of common genetic risk on ADHD diagnosis or symptoms (Hermosillo et al. 2020; Mooney et al. 2021; Sudre et al. 2020). A reasonable question is whether a brain-wide summary score will mediate a greater proportion of polygenic risk than individual brain features.

Our goals for this study were: (a) to develop a PNS associated with ADHD symptoms, (b) to examine whether brain networks/connections that contribute most to this ADHD PNS are consistent with previous rs-fcMRI studies (i.e., default mode and cognitive control networks), (c) to validate the predictive ability of the ADHD PNS in a completely independent longitudinal ADHD case-control cohort, and (d) to test whether the ADHD PNS mediates common genetic risk for ADHD, or whether effects are additive.

This work stands to provide evidence for the validity of brain-wide summary measures from neuroimaging data for the prediction of behavior traits, and to demonstrate how these scores can be used to provide insight into mechanistic and causal pathways.

## MATERIALS AND METHODS

### Participants

We report on data from two independent cohorts. The Adolescent Brain Cognitive Development (ABCD) study served as our discovery cohort (Casey et al. 2018). It is a 21-site, diverse cohort of over 11,000 children (age 9-10 at baseline). The children have been genotyped and are followed with extensive behavioral, cognitive, clinical, and MRI measures annually. The data set is publicly available on the NIMH Data Archive (http://dx.doi.org/10.15154/1504041) (Feczko et al. 2021).

For internal replication, we split the ABCD cohort into two ABCD Reproducible Matched Samples (ARMS) matched on nine demographic factors thought to be involved with development: site, age, assigned sex at birth, race/ethnicity, grade, highest level of parental education, handedness, combined family income, and exposure to anesthesia (Feczko, Earl, Perrone, and Fair 2020b; 2020a).

The Oregon-ADHD-1000 cohort (Karalunas et al. 2017; Nigg et al. 2018; Nigg, Karalunas, Gustafsson, et al. 2020; Mooney et al. 2020; 2021) was used as an independent validation data set. It is a case-control cohort of ∼1400 children, age 7-11 years at baseline, that is enriched for psychopathology by virtue of requiring ∼2/3rds of the sample to meet detailed research and DSM criteria for ADHD at baseline. Of these, 553 have computational measures of umbrella executive functioning (working memory, response inhibition, arousal regulation, reward discounting), clinical measures, and MRI data at one or more time points. Genome-wide genotype data was available for 487 of these children (NDA Collection 2857).

### Resting-state functional connectivity MRI data

Minimally processed, quality-controlled, baseline resting-state functional MRI data for the ABCD cohort were downloaded from the NIMH Data Archive (NDA), specifically the ABCD-BIDS Community Collection (ABCC; NDA Collection 3165). Details of the data access, download, and processing can be found in the ABCC documentation (https://collection3165.readthedocs.io/en/stable/). Briefly, the data was processed using the ABCD-BIDS pipeline (Feczko, Earl, Perrone, and Fair 2020c), which is a modified version of the Human Connectome Project (HCP) pipeline (Feczko et al. 2021; Glasser et al. 2013). Processed ABCD data were further curated based on head motion, such that only frames that had a framewise displacement (FD) threshold of 0.2mm were used. Of the surviving frames, additional frames were removed if they were detected as outliers (using the median absolute deviation) in the standard deviation of the bold signal across the whole brain. Trimming was done at random to limit bias in data selection. Only resting state scans from participants trimmed to 10 minutes of usable data were kept for further analysis. The final ABCD data set for our primary analyses consisted of N=2801 subjects in ABCD ARMS-1 and N=2865 subjects in ABCD ARMS-2. Secondarily, to ensure findings were robust to the effects of motion during scan acquisition, a sensitivity analysis was performed on a subset of subjects (combined ABCD ARMS, N=2936) using a stricter FD threshold (0.1mm). Results were compared for analyses using 8 minutes of data at an FD threshold of 0.2mm vs. an FD threshold of 0.1mm.

Whole-brain functional connectivity was measured using the Gordon parcellation, an independent annotation derived from highly sampled individual data, which includes subcortical nuclei and the cerebellum (Gordon et al. 2016).

The Oregon cohort rs-fcMRI data was processed using the same pipeline as the ABCD cohort. After quality control (poor image quality, registration, or grey matter/white matter delineation) and exclusion of subjects with missing data in the Oregon cohort, a total of N=494 subjects were kept for the baseline analysis, and N=553 for longitudinal analyses.

### ADHD composite symptom scores

ADHD symptom scores were created by averaging standardized (Z-scored) symptom measures across available measures in both the ABCD and Oregon-ADHD-1000 cohorts. For ABCD, the measures used were the ADHD symptom count from the Child Behavior Checklist (CBCL), and the inattentive and hyperactive symptom counts from the Kiddie Schedule for Affective Disorders and Schizophrenia (KSADS). This parent-reported composite score was previously validated and replicated for ABCD in a confirmatory factor model with reproducible fit (Cordova et al. 2021). For the Oregon-ADHD-1000, the measures used to create a composite were the inattentive and hyperactive symptom counts from the ADHD Rating Scale, the KSADS, the Conners (3^rd^ edition), and the Strengths and Weaknesses of Attention-Deficit/Hyperactivity Disorder Symptoms and Normal Behavior Scale (SWAN), as well as the hyperactivity raw score from the Strengths and Difficulties Questionnaire (SDQ) (Nigg, Karalunas, Gustafsson, et al. 2020).

### Brain-wide association analyses

Analyses testing the association between brain connectivity (i.e., each parcel-to-parcel brain correlation) and the ADHD composite symptom scores in ABCD were carried out using the Sandwich Estimator for Neuroimaging Data (SEND) (Feczko et al. 2021; Feczko, Earl, Perrone, Fair, et al. 2020). SEND constructs a marginal model (Guillaume et al. 2014) for brain-wide associations, where brain connectivity is the dependent variable, while covariates and independent variables are modeled as fixed effects. Standard errors are estimated with a Sandwich Estimator clustered for unique combinations of race, ethnicity, study site, and gender. Two covariates, subject age and highest parental education, and the independent variable ADHD composite symptom scores were modeled as fixed effects.

### Brain-wide connectivity (polyneuro) scores

Polyneuro scores (PNS), which represent brain-wide connectivity associated with ADHD symptoms, were created following a method (Figure 1) similar to that described previously (Zhao et al. 2019). Effect estimates generated from the brain-wide association study performed on the ABCD discovery set were used to calculate PNS for each subject in the validation data sets by multiplying the effect estimate for each connection, *β*_*i*_, by the subject’s connectivity measure, *C*_*i*_, and summing across all connections (Equation 1). Scores were created using both unadjusted effect estimates, as well as effect estimates adjusted using a Bayesian procedure described previously (Zhao et al. 2019). The Bayesian adjustment accounts for both the observed correlation structure among all connections brain-wide, as well as the signal-to-noise ratio of the brain-behavior association. In addition, scores were created using only those connections that passed a particular significance threshold: top 50%, top 10%, and top 1% most significant connections.

**Figure 1.**
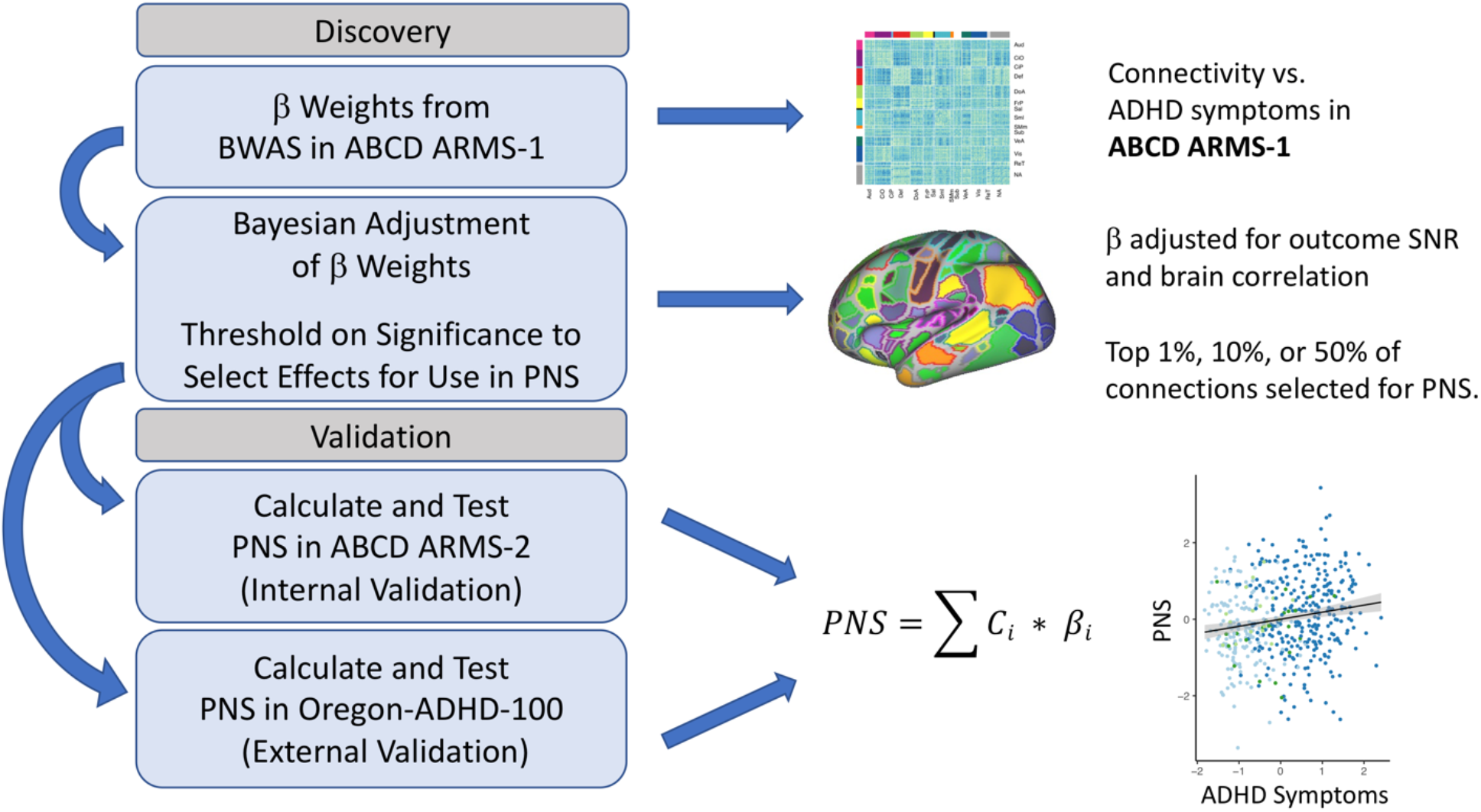
Polyneuro score (PNS) discovery and validation workflow.

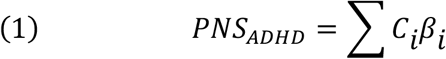

The association between the PNS and ADHD composite symptoms score was validated in both the ABCD cohort and the completely independent Oregon-ADHD-1000 cohort. First, ABCD ARMS-1 was used as the discovery cohort, and effect estimates were used to calculate PNS in ABCD ARMS-2. The association between ADHD PNS and ADHD symptoms was tested using a linear regression model, with age, sex, race/ethnicity, highest parental education, ABCD study site, and the single most significant connection covaried. To assess whether family relatedness impacted the PNS effect estimate, marginal models with family ID as the cluster variable were also fit. Marginal models were implemented with the gee R package (Carey 2019).

Second, the full ABCD cohort (ARMS-1 and ARMS-2 combined) was used as the discovery data set, and ADHD PNS scores were calculated for all subjects in the Oregon-ADHD-1000 cohort. The association between ADHD PNS and ADHD composite symptom score was validated in the Oregon cohort in three ways: (1) The earliest scan was chosen for each subject and the association between PNS and ADHD symptoms was tested in this baseline data using linear regression. Again, a marginal model with clusters defined by family ID was fit to verify effect estimates while accounting for the impact of siblings. (2) All available scans were used and repeated measures were accounted for with a marginal model, again using the gee R package. (3) Finally, subjects were categorized as follows: persistent ADHD (ADHD at all time points), persistent control (control at all time points), remittent ADHD (ADHD at one or more early time points followed by non-ADHD status), and other. Again, a marginal model was fit to assess the association between these diagnostic categories and the ADHD PNS over time. All models fit in the Oregon cohort included age, assigned sex at birth, race/ethnicity, highest parental education, puberty score, scan type, and the most significant individual connection as covariates.

### ADHD Polygenic Score

Details of the Oregon-ADHD-1000 genotyping procedure have been published previously (Nigg et al. 2018; Nigg, Karalunas, Gustafsson, et al. 2020; Mooney et al. 2021). Briefly, DNA was extracted from saliva collected in Oragene(R) cups (DNA Genotek Inc., Kanata, ON, Canada) using manufacturer’s protocols. All subjects were genotyped at the Stanley Center for Psychiatric Research (Broad Institute of MIT and Harvard, Cambridge, MA) using the Illumina PsychCHIP v1-1.

For the ABCD cohort, saliva samples were collected at the baseline visit and sent to the Rutgers University Cell and DNA Repository for DNA isolation (Uban et al. 2018). Genotyping was performed using the Smokescreen Array (Baurley et al. 2016). For the present study, processed genotypes were downloaded from the NIMH Data Archive (dx.doi.org/10.15154/1503209), and standard QC checks were performed. One batch of samples had significantly lower call rate (∼85%) than all others (∼98%) and was removed (N=126 samples). All SNPs had adequate call rate (>94%), and no significant batch effects were observed after examining allele frequency differences across batches.

For both cohorts, principal component analysis was conducted using the PC-Air method in the GENESIS Bioconductor package (Gogarten et al. 2019), and nongenotyped SNPs were imputed with IMPUTE2 (Howie, Donnelly, and Marchini 2009) using the 1000 Genomes phase 3 reference panel. Only those SNPs imputed with high confidence (INFO > 0.8) were kept. Genotype probabilities were converted to best-guess genotypes, with the genotype set to missing if the probability < 0.8.

The polygenic risk scores (PRS) were constructed in both ABCD and Oregon-ADHD-1000 using the 2016-2017 PGC+iPSYCH ADHD GWAS meta-analysis (Demontis et al. 2019) as the discovery data set (20183 ADHD cases; 35191 controls). The PRS was calculated using the LDpred method (Vilhjálmsson et al. 2015). Only SNPs with INFO score > 0.8 in both the PGC+iPSYCH meta-analysis and the target data sets were considered. SNPs were further limited to the ∼1.2 million HapMap SNPs as suggested. Linkage disequilibrium was estimated using all unrelated individuals in the ABCD cohort, and PRS were created with the proportion of causal SNPs set to 0.3.

Given that European-ancestry individuals are the largest ancestry group in both the ABCD and Oregon cohorts, we also examined an ADHD PRS constructed using the European-only PGC+iPSYCH meta-analysis (Demontis et al. 2019) as the discovery data set (19099 ADHD cases and 34194 controls). Other than the discovery data used, methods for constructing the PRS were the same as above.

In all analyses examining the effect of the ADHD PRS, the first three genomic principal components were covaried to adjust for potential population stratification. Analyses examining whether the effect of the PRS on ADHD symptoms is partially mediated by brain-wide connectivity (ADHD PNS) were conducted using the mediation R package (Tingley et al. 2014).

## RESULTS

### Overview of samples

There were no differences in mean age or race/ethnicity between the two ABCD ARMS, but a slight difference in the proportion of females (Chi-square p-value=0.0463) (Table 1). The distribution of symptom scores was not significantly different between ABCD ARMS-1 and ARMS-2 (p=0.916).

**Table 1.** Description of Cohorts

|  | ABCD ARMS-1 | ABCD ARMS-2 | Oregon-ADHD-1000 |
| --- | --- | --- | --- |
| Sample Size | 2801 | 2865 | 553 |
| Mean Age (SD) | 9.97 (0.63) | 9.98 (0.62) | 10.25 (1.56)* |
| % Female | 52.9 | 50.2 | 37.4 |
| % Caucasian | 65.0% | 65.5% | 79.7% |

\* Age at earliest scan.

By virtue of its case-control design, the Oregon-ADHD-1000 had a higher proportion of ADHD cases and as a result, much greater variation in symptom scores compared to ABCD (Figure S1). The Oregon cohort also had a lower proportion of females and a greater proportion of Caucasians than the ABCD cohort (p-values<0.001) (Table 1).

### Brain-wide association results

Brain-wide association analyses were performed in ABCD ARMS-1 and ARMS-2 separately, as well as in the full ABCD cohort. Correlation of effect estimates between ARMS-1 and ARMS-2 was significant, with Pearson’s correlation ranging between 0.46 for all connections and 0.79 for the top 1% most significantly associated connections (Table 2).

**Table 2.** Correlation of Brain-wide Effect Estimates Across ABCD ARMS

| T-stat Quantile | Pearson’s Correlation | P-value |
| --- | --- | --- |
| All | 0.460 | 0 |
| Top 50% | 0.571 | 0 |
| Top 10% | 0.717 | 0 |
| Top 1% | 0.792 | 0 |

Effect estimates for individual connections were small, with standardized regression coefficients <0.1. A matrix of regression coefficients for all connections brain-wide is shown in Figure 2A. The top 10% most significant associations are spread across multiple brain networks, but are particularly concentrated among connections involving the default mode and cingulo-opercular networks (Figure S2). The cumulative effect of connections involving each ROI (i.e., the sum of the absolute value of effect estimates across each row in the matrix), along with each ROI’s network assignment, are shown in Figure 2B.

**Figure 2.**
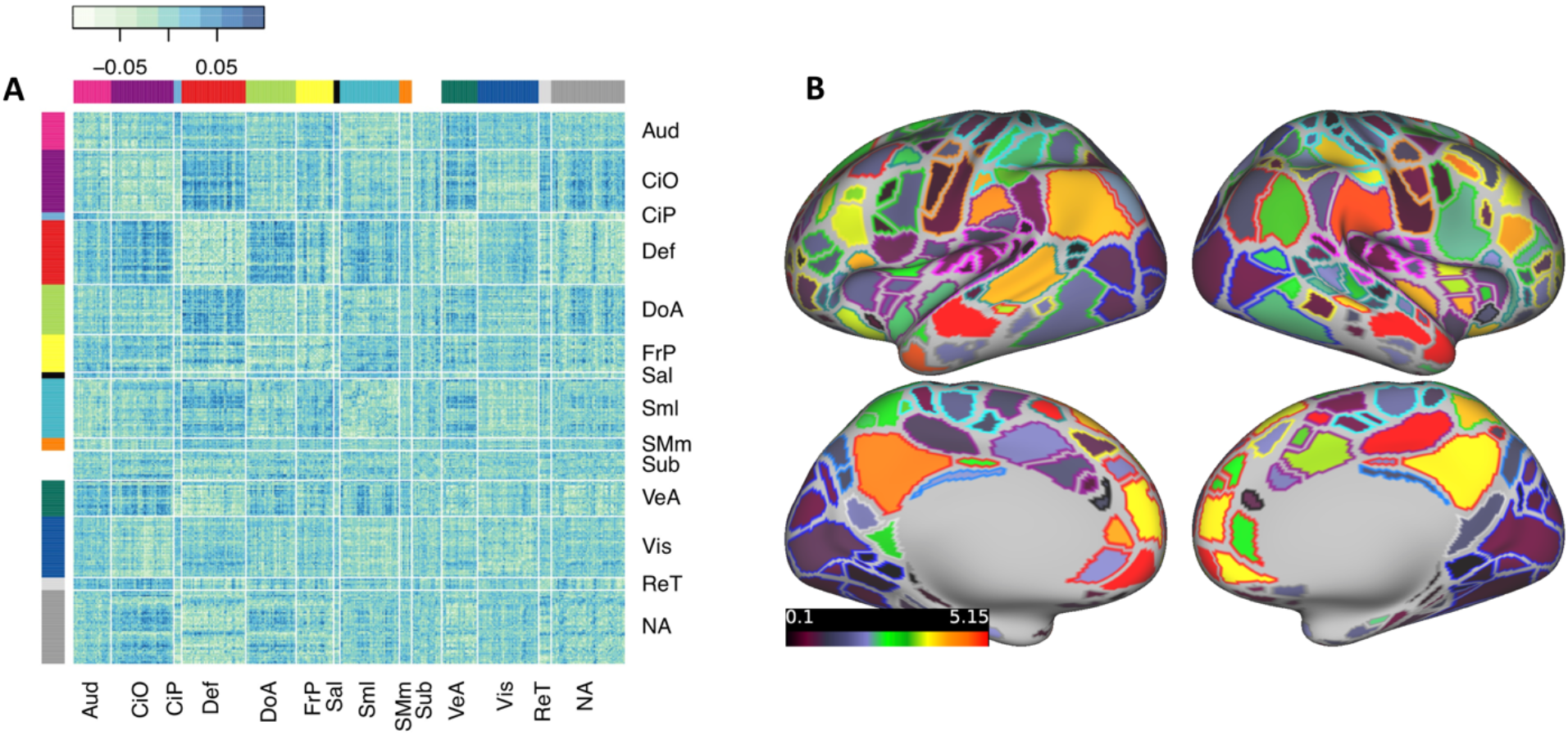
Brain-wide connectivity associated with ADHD symptoms in the ABCD study cohort. **(A)** The matrix of standardized regression coefficients showing the strength of association between all connections (organized by brain network) and ADHD symptoms. **(B)** Gordon parcellation showing the relative contribution of each brain network to the ADHD PNS. Only the top 10% most significant connections (representing the most predictive PNS) are considered. Fill color represents the sum of the absolute value of beta weights for all connections in which a parcel participates; outline color represents network assignment. Aud = Auditory, CiO = Cingulo-opercular, CiP = Cingulo-parietal, Def = Default Mode, DoA = Dorsal Attention, FrP = Fronto-parietal, Sal = Salience, Sml = Somato-sensory Hand, SMm = Somato-sensory Mouth, Sub = Subcortical, VeA = Ventral Attention, Vis = Visual, ReT = Retrosplenial Temporal, NA = Not Assigned.

### Polyneuro scores associated with ADHD symptoms

To assess whether the cumulative effect of brain-wide, ADHD-associated resting-state connectivity could predict ADHD symptoms, PNS were constructed based on the effect estimates from BWAS in ABCD ARMS-1 and the full ABCD cohort.

The ADHD PNS (Bayesian-adjusted, top 10% of connections), based on effect estimates discovered in ABCD ARMS-1, was significantly associated with ADHD symptom scores in ABCD ARMS-2 (β=0.078 (0.033, 0.123); p=7.4e-4; Figure 3A), after adjusting for the single most significant connection. Not surprisingly, given the small number of ADHD cases present in ABCD (Figure S1), the PNS explained a very small proportion of symptom variation (partial R^2^=0.004).

**Figure 3.**
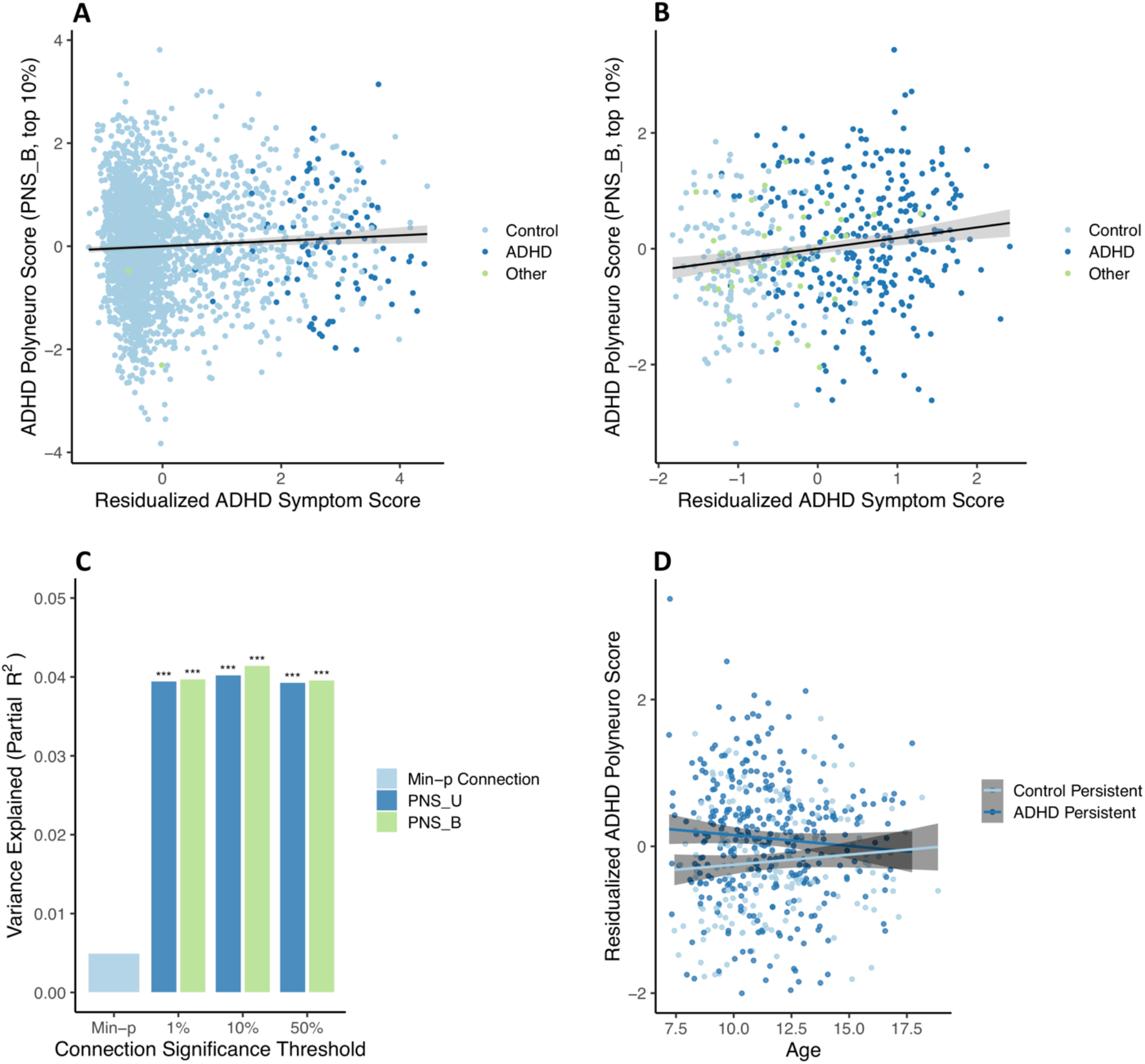
ADHD polyneuro score associated with ADHD symptoms. Polyneuro scores and residualized ADHD symptom scores, after adjusting for relevant covariates (see Methods), for all subjects in the **(A)** ABCD ARMS-2 (N=2865) and **(B)** Oregon-ADHD-1000 cohort, using each subject’s earliest scan (N=494). **(C)** The proportion of ADHD symptom score variance explained by the single most significantly associated connection, polyneuro scores comprised of the top 1%, 10%, and 50% most significant connections in the Oregon cohort. **(D)** Subjects in the Oregon-ADHD-1000 cohort with persistent ADHD showed higher ADHD PNS than controls (p=0.00142), though this difference decreases with age. PNS_U = unadjusted polyneuro score, PNS_B = Bayesian adjusted polyneuro score. *** = p < 0.001.

The full ABCD sample was used as the discovery cohort for validating the ADHD PNS in the Oregon-ADHD-1000 cohort. When examining baseline data only (i.e., the earliest scan for each subject in the Oregon cohort; N=494), the ADHD PNS was significantly associated with ADHD symptoms, after accounting for relevant covariates (see Methods) and the single most significantly associated connection (Figure 3B). The Bayesian-adjusted PNS based on the top 10% most significant connections was the most predictive (β=0.208 (0.113, 0.302); p=2.1e-5), explaining approximately 4.1% of the variation in the ADHD composite symptom score. The amount of variance explained by the PNS is roughly 8 times that explained by the single most significant connection (∼0.5%), demonstrating the benefit of the brain-wide score (Figure 3C). Although the Bayesian-adjusted PNS were slightly more predictive than the unadjusted PNS (Figure 3C), this difference was not significant (all p-values > 0.05).

Results are confirmed (β=0.221 (0.119, 0.322); p=2.5e-5) when examining only subjects <12 years of age (N=416), to better match the ABCD discovery cohort. Furthermore, the effect estimate from a marginal model, with clusters defined by family ID, is nearly identical (β=0.208, robust p=2.3e-6), suggesting family structure has minimal influence on the associations observed.

Including all scans in the analysis (N=553 subjects, N=1048 scans) and using a marginal model to account for repeated measures of the same individual resulted in a much reduced, but still significant, association between the PNS and ADHD symptoms (β=0.0626 (0.016, 0.110), robust p=0.00903). This smaller effect is consistent with the strength of association between the PNS and ADHD symptoms decreasing with age (see below).

Examining group effects among youth in the Oregon ADHD-1000 with at least two scans (N=320 subjects, N=806 scans) showed that subjects with persistent ADHD had significantly higher PNS than healthy controls across time (control group β=-0.331 (−0.518, -0.143), robust p=5.5e-4) (Figure 3D). There was a borderline significant interaction between ADHD status and age at scan (interaction β=0.0585 (−0.002, 0.119), robust p=0.060), suggesting the difference in PNS between ADHD cases and controls diminishes with increasing age (Figure 3D).

To ensure that findings were not significantly impacted by, or attributed to, motion during scan acquisition, we repeated analyses while accounting for motion in two ways. First, using a subsample of the ABCD discovery cohort (N=2936), BWAS were conducted using two FD thresholds (0.1mm vs. 0.2mm; 8 minutes of data for both), and results from each were used to construct PNS in the Oregon cohort. The two resulting PNS had nearly identical predictive power (Figure S3), suggesting motion in the discovery cohort did not significantly impact the PNS.

Second, an estimate of motion during each scan (mean FD) was included in regression models when testing the association between PNS and ADHD symptoms in the Oregon cohort. This estimate of motion during scan acquisition was not significantly associated with ADHD symptoms (p=0.690 for the baseline analysis), and did not meaningfully change the strength of association between symptoms and the PNS (β=0.202 after adjusting for mean FD in the baseline analysis).

### ADHD polyneuro score relationship to ADHD polygenic risk score

In the ACBD ARMS-2, there was a weak but significant correlation between the PNS and ADHD PRS (r=0.177, p<2e-16). However, when included in the same regression model, both the ADHD PNS (β=0.071 (0.024, 0.118), p=0.0032) and PRS (β=0.178 (0.132, 0.225), p=4.6e-14) were significantly associated with symptoms, and there was no significant mediation of the genetic effect by the PNS (bootstrapped p=0.11). The PRS effect was stronger for the score based on the European-ancestry discovery GWAS (β=0.231 (0.168, 0.294), p=6.7e-13), and a small, borderline significant mediation effect was observed (bootstrapped p=0.052, proportion mediated=0.016).

In the Oregon cohort, similarly, both the ADHD PNS (β=0.217 (0.112, 0.322); p=6.3e-5) and the ADHD PRS (β=0.215 (0.125, 0.306); p=4.1e-6) were significantly associated with ADHD symptoms when included in the same regression model. However, the two scores were not correlated (p=0.857), and a mediation model showed no indication of the PNS mediating genetic risk (bootstrapped p=0.90). As in ABCD ARMS-2, the PRS effect was slightly stronger when using the European-ancestry discovery data (β=0.232 (0.135, 0.329); p=3.7e-6), but still no mediation was seen (bootstrapped p=0.91)

## DISCUSSION

### PNS Reveals a Distributed View of Brain-wide Association with ADHD Symptoms

Our findings demonstrate a robust association between brain-wide connectivity patterns (PNS) and ADHD symptoms in two independent cohorts with different recruitment procedures, supporting generalizability of these findings. PNS that represent cumulative, brain-wide effects explained a significantly greater proportion of the variation in symptom scores than the most significant individual connections. Although the strongest connectivity-symptom associations that contribute to the PNS were concentrated within connections involving the default mode and cingulo-opercular networks, the most predictive PNS incorporated connections involving all networks (Figure S3), and explained ∼4% of the variation in symptoms—roughly 8 times the variation explained by the single most significant connection. In the Oregon cohort, subjects with an ADHD PNS in the highest 10% had 3.5 times greater odds of an ADHD diagnosis than those with a PNS in the lowest 10%, demonstrating the potential predictive utility of the brain-wide summary measure. The explanatory power of the PNS examined here is comparable to polygenic risk scores and will likely improve with larger discovery data sets (i.e., more precise effect estimates for individual connections)—likewise, the specific network contributions to the brain-wide score should be further evaluated and clarified with additional data.

Our results are consistent with previous results implicating the default mode and cognitive controls networks in ADHD, but also demonstrate that focusing on the effects of individual networks does not fully capture resting-state connectivity patterns associated with the disorder. Such results support recent reports of distributed, brain-wide associations with other cognitive traits. In the context of the recent literature, our findings emphasize a distributed view of brain function, where complex cognitive behaviors or traits are best explained by cumulative effects across most brain networks (Zhao et al. 2019; Marek et al. 2020).

### ROI-based PNS Were Unaffected by Correlations between Connections

Sensitivity analyses were conducted to ensure that potential sources of bias (relatedness among subjects and motion during MRI acquisition) were accounted for, lending confidence to our results. In addition, while Bayesian-adjusted PNS, which account for outcome signal-to-noise ratio and the correlation of effects across the brain, were slightly more predictive than unadjusted PNS, the difference was not significant, contrary to previous work (Zhao et al. 2019). It is possible that our use of parcellated connectivity data, which already accounts for spatial correlation across the brain, reduced the need for this adjustment.

### Lack of Genetic Risk Mediation Suggests Role for Environmental/Developmental Context in ADHD Symptoms

ADHD PNS did not mediate common genetic risk for ADHD (PRS), suggesting that future research should investigate environmental factors associated with ADHD-related connectivity patterns. It also suggests the assumption that genetic effects on ADHD are mediated principally through neural networks may be insufficient. This is consistent with low to moderate heritability for many brain features (Adhikari et al. 2018; Miranda-Dominguez et al. 2017; Sudre et al. 2017; Miranda-Dominguez et al. 2017) and the growing recognition of the important role of environmental and developmental context in combination with genetic liability in ADHD. More work is needed to disentangle environmental confounds from such mediation analyses.

We used a method for calculating polygenic scores that has demonstrated good performance in a variety of data sets, as well as significantly improved performance compared to more traditional methods (Vilhjálmsson et al. 2015; Ni et al. 2021; Pain et al. 2021). We also constructed PRS based on both the full and European-ancestry-only PGC+iPSYCH ADHD GWAS meta-analyses, to examine whether the discovery sample used significantly impacted our results. While we did see small differences in PRS effects between the two discovery sets, mediation results did not change meaningfully. However, it is important to note that larger samples size, both in terms of discovery (for PNS and PRS) and validation, may allow for more precise scores and the detection of small but meaningful mediation effects. In addition, access to more diverse discovery cohorts for PRS are sorely needed to address the generalizability of PRS effects (Martin et al. 2019).

### Longitudinal Extension of PNS Models Can Assess Specificity to Childhood ADHD

Another important direction for future work should be the examination of how ADHD-associated connectivity changes with age (Kessler, Angstadt, and Sripada 2016). The PNS reported here were derived from data on children 9-10 years of age in the ABCD sample, and showed at least suggestive evidence in the Oregon cohort of reduced predictive power in older subjects. This result raises the question of whether the scores examined here are specific to childhood ADHD (i.e., that older discovery cohorts should be used for predicting in older populations), or whether brain connectivity is simply less predictive of ADHD in older individuals. Subsequent waves of data released from the ABCD study will allow further exploration of age effects.

### PNS Approach Leverages Small Effect Sizes to Develop Generalizable Brain Measures for Clinical Research

Overall, the findings reported here provide strong evidence for the use of brain-wide summary measures of resting-state connectivity for predicting ADHD. These polyneuro scores have several advantages, beyond the fact that they are significantly more predictive than individual brain features. First, because of the larger effect sizes for PNS, they will increase the utility of small neuroimaging data sets that are underpowered to detect associations with individual brain features. Secondly, PNS partially alleviate issues of heterogeneity, given that the same cumulative risk can be the result of different individual risk factors. Third, PNS may provide new ways to examine shared brain mechanisms across disorders (e.g., does an ADHD PNS predict depression symptoms?), and provide insights into the relationships between diagnostic categories and the risk of psychiatric comorbidities. And finally, further development of the PNS approach and integration with other risk factors (e.g., polygenic and environmental measures) has the potential to provide clinically meaningful prediction of behavioral disorders and patient outcomes.

## Data Availability

All data used in the present study is publicly available on the NIMH Data Archive.

http://dx.doi.org/10.15154/1504041

https://collection3165.readthedocs.io/en/stable/

https://nda.nih.gov/edit_collection.html?id=3165

https://nda.nih.gov/edit_collection.html?id=2857

## ACKNOWLEDGEMENTS

This work was supported by funding from the National Institute of Mental Health of the National Institutes of Health under award numbers: R01MH115357 (Fair and Nigg), R37MH059105 (Nigg), U01DA041148 (Fair, Nagel, Feldstein Ewing), and U24DA041123 (Fair). The content is solely the responsibility of the authors and does not necessarily represent the official views of the NIH. Creation of genetic scores benefited from the collaboration of the OHSU Integrated Genomics Core (Chris Harrington, Ph.D., Director) and Kristina Vartanian.

Large-scale image processing across the data set greatly benefited from collaboration with the Masonic Institute for the Developing Brain Informatics Group (MIDB-IG, Thomas Pengo, Ben Lynch, Timothy Hendrickson, James Wilgenbusch).

## SUPPLEMENTAL FIGURES

**Figure S1.**
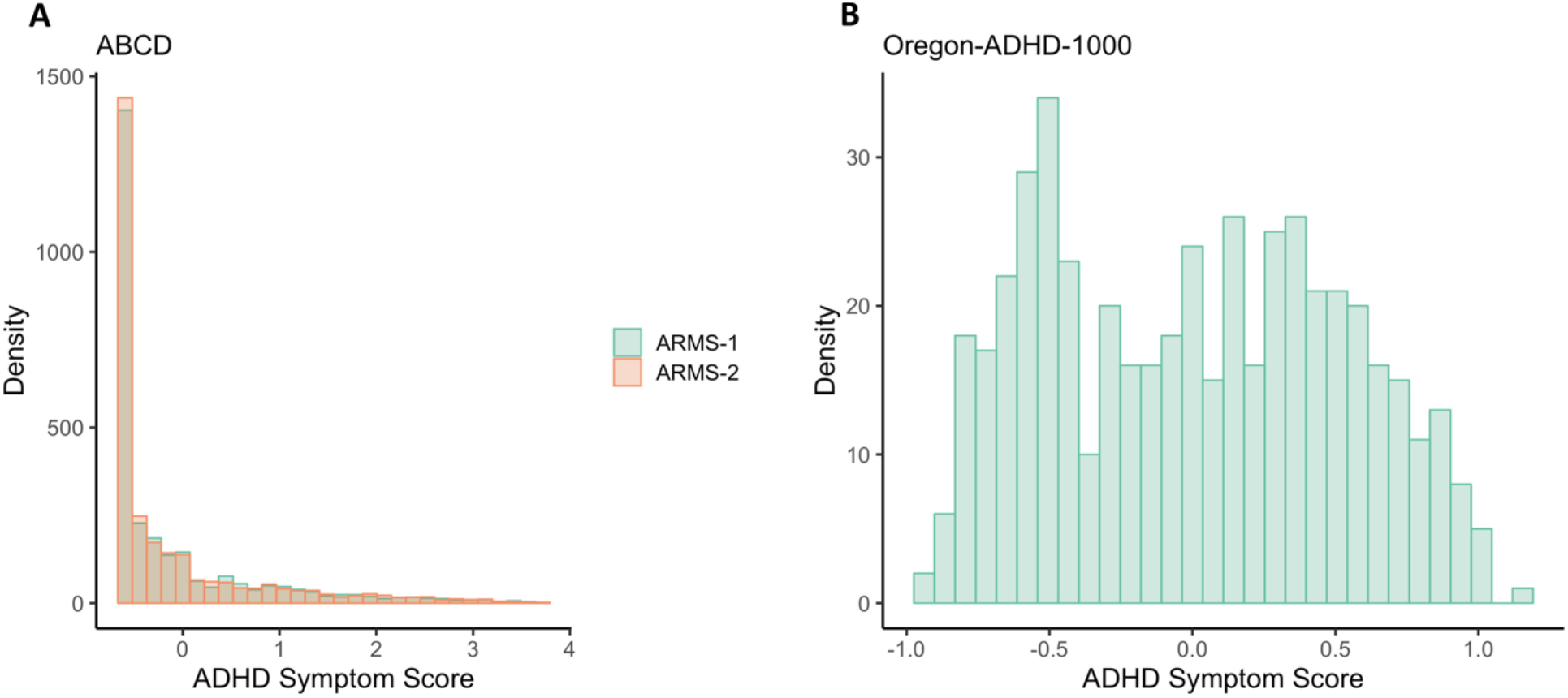
Distribution of ADHD symptom scores in **(A)** both ARMS of ABCD (Mann-Whitney test p-value=0.916), and **(B)** the Oregon-ADHD-1000 case-control cohort. The ADHD composite symptom scores are the average of multiple standardized (mean=0, SD=1) symptom scales (see Methods).

**Figure S2.**
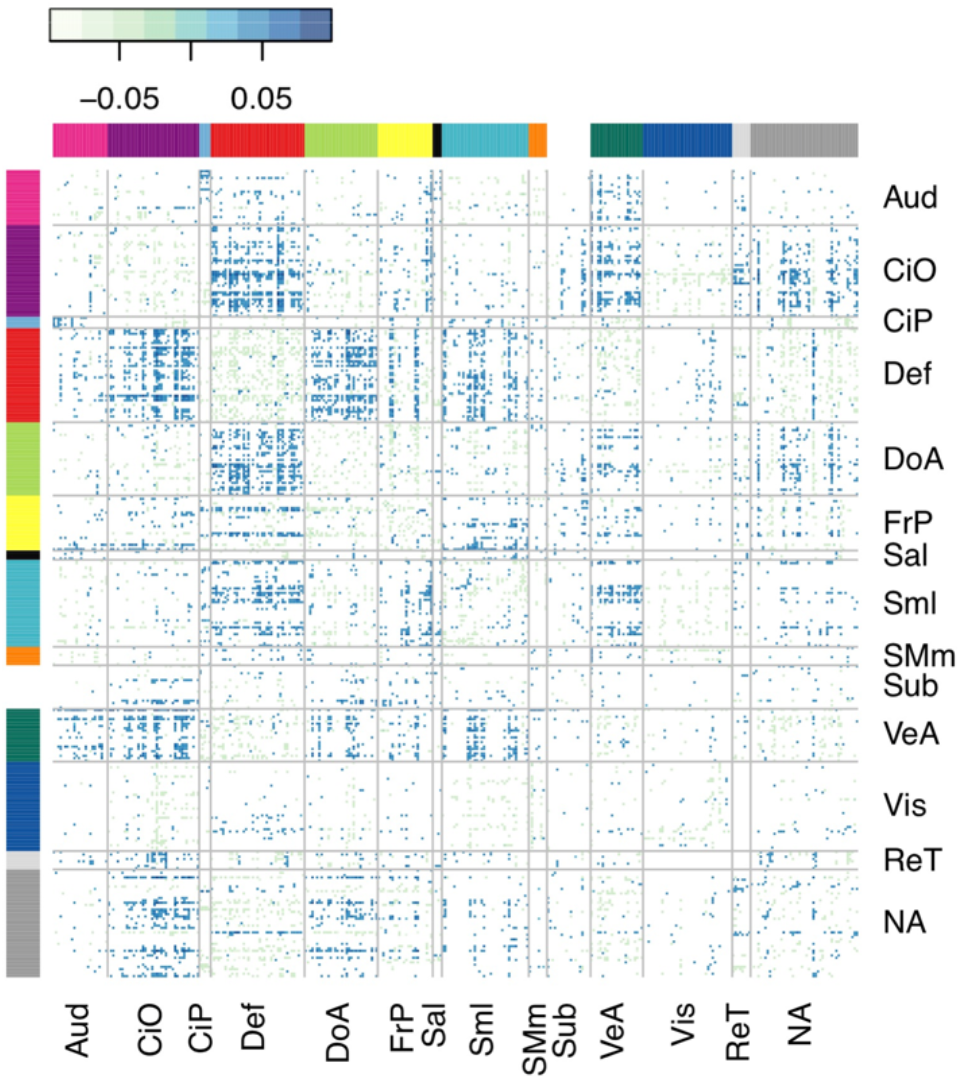
The matrix of standardized regression coefficients showing the strength of association between the top 10% most significant connections (organized by brain network) and ADHD symptoms. The cumulative effect of these connections comprised the most predictive PNS, demonstrating the brain-wide, distributed nature of the ADHD PNS. Aud = Auditory, CiO = Cingulo-opercular, CiP = Cingulo-parietal, Def = Default Mode, DoA = Dorsal Attention, FrP = Fronto-parietal, Sal = Salience, Sml = Somato-sensory Hand, SMm = Somato-sensory Mouth, Sub = Subcortical, VeA = Ventral Attention, Vis = Visual, ReT = Retrosplenial Temporal, NA = Not Assigned.

**Figure S3.**
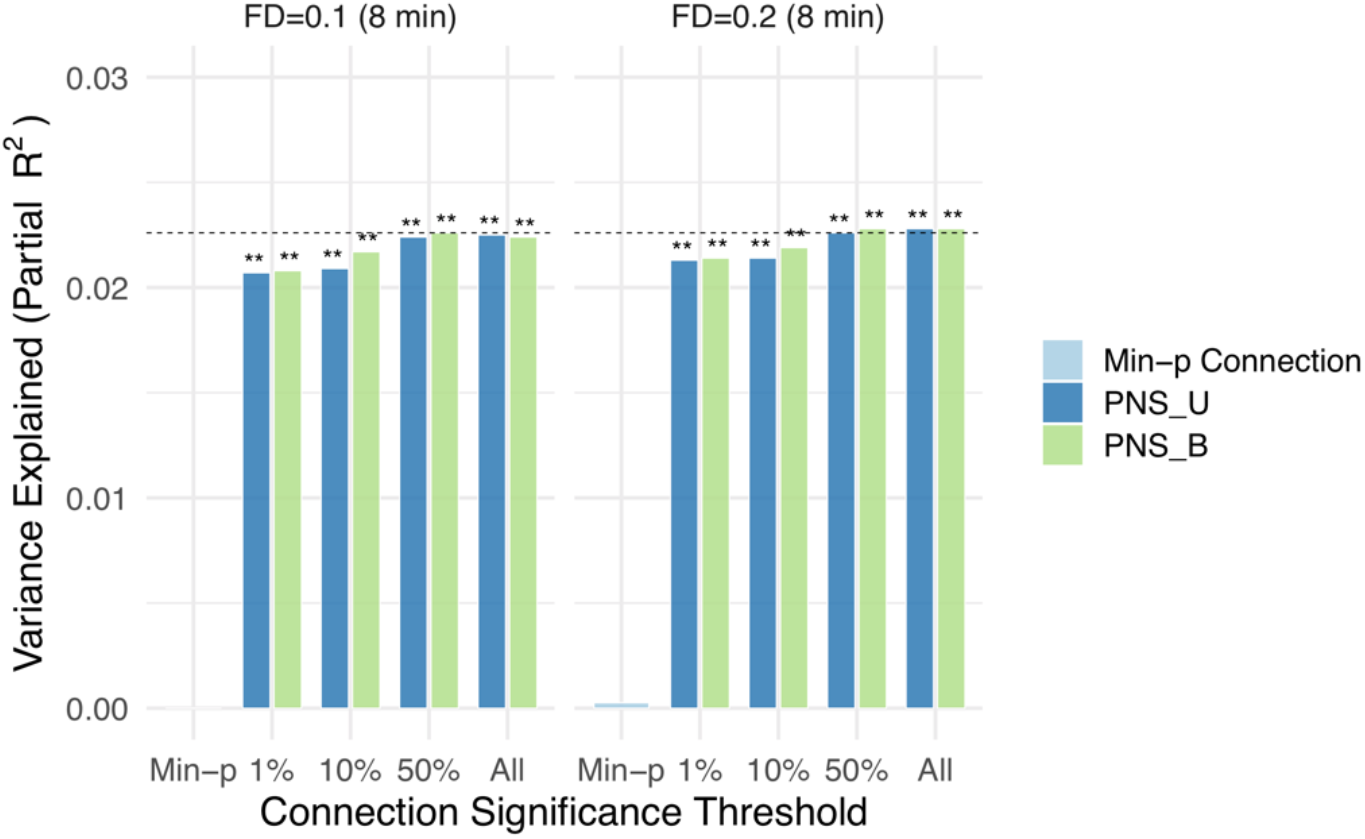
ADHD polyneuro scores were robust to motion threshold. The proportion of ADHD symptom score variance explained is nearly identical when analyzing the same set of subjects (N=2936) using a more stringent motion threshold (framewise displacement threshold of 0.1mm vs. 0.2mm). PNS_U = unadjusted polyneuro score, PNS_B = Bayesian adjusted polyneuro score. ** = p < 0.01.

